# Introduction and Establishment of SARS-CoV-2 Gamma Variant in New York City in Early 2021

**DOI:** 10.1101/2022.04.15.22273909

**Authors:** Tetyana I. Vasylyeva, Courtney E. Fang, Michelle Su, Jennifer L. Havens, Edyth Parker, Jade C. Wang, Mark Zeller, Anna Yakovleva, Gabriel W. Hassler, Moinuddin A. Chowdhury, Kristian G. Andersen, Scott Hughes, Joel O. Wertheim

**Author notes:** To whom correspondence should be addressed: Tetyana I. Vasylyeva.

## Abstract

**Background:** Monitoring the emergence and spread of SARS-CoV-2 variants is an important public health objective. Travel restrictions, aimed to prevent viral spread, have major economic consequences and unclear effectiveness despite considerable research. We investigated the introduction and establishment of the Gamma variant in New York City (NYC) in 2021.

**Methods:** We performed phylogeographic analysis on 15,967 Gamma sequences available on GISAID and sampled between March 10^th^ through May 1^st^, 2021, to identify geographic sources of Gamma lineages introduced into NYC. We identified locally circulating Gamma transmission clusters and inferred the timing of their establishment in NYC.

**Findings:** We identified 16 phylogenetically-distinct Gamma clusters established in NYC (cluster sizes ranged 2-108 genomes). Most of the NYC clusters were introduced from Florida and Illinois; only one was introduced from outside the United States (US). By the time the first Gamma case was reported by genomic surveillance in NYC on March 10^th^, the majority (57%) of circulating Gamma lineages had already been established in the city for at least two weeks.

**Interpretation:** Despite the expansion of SARS-CoV-2 genomic surveillance in NYC, there was a substantial gap between Gamma variant introduction and establishment in January/February 2021, and its identification by genomic surveillance in March 2021. Although travel from Brazil to the US was restricted from May 2020 through the end of the study period, this restriction did not prevent Gamma from becoming established in NYC as most introductions occurred from domestic locations.

## BACKGROUND

Monitoring the emergence, introduction, and circulation of severe acute respiratory syndrome coronavirus 2 (SARS-CoV-2) Variants of Concern (VOCs) and Variants of Interest (VOIs) is an important public health tool that is essential to guiding an effective pandemic response (1). Emergence and rapid spread of SARS-COV-2 variants has driven consecutive epidemic waves regionally and around the globe, starting with the Alpha variant (B.1.1.7 lineage) identified in the UK in late 2020, followed by the Delta variant (B.1.617.2) identified in India in early 2021, and the current wave of Omicron variant infections first identified in South Africa in late 2021. In the US and elsewhere, the response to the identification of these variants has included restricting travel from locations where the variants were first identified; however, widespread establishment of these variants nonetheless ensued.

Gamma VOC (P.1 lineage) was first identified in late 2020 in Manaus, Brazil, a region that previously experienced a high COVID-19 burden and had an estimated 68% seroprevalence (2). The rapid spread of Gamma in a region with high pre-existing immunity immediately raised concerns about the transmission potential of this variant. On January 25^th^, 2021, the first case of Gamma infection was documented in the United States (US) in the state of Minnesota (3). Travel from Brazil to the US was prohibited for non-US citizens starting May 2020, and this was extended indefinitely on January 26^th^, 2021 due to concern about Gamma spread (4). Nevertheless, by May 2021, Gamma had been detected in at least 41 US states (https://outbreak.info/).

Molecular epidemiology investigations into this lineage characterized 17 novel mutations relative to the wild type virus (Hu-1 reference strain)—including ten mutations in the spike protein, three of which (K417T, E484K, and N501Y) have been shown to increase spike ACE2 receptor binding affinity (5)—leading to the lineage being designated a VOC by the CDC (6). Gamma virus is poorly inhibited by convalescent plasma and sera from vaccinated individuals (7), is partially resistant to monoclonal antibodies used for COVID-19 treatment (8), and might be associated with increased COVID-19 severity and mortality in younger adults and adults without comorbidities, compared to the wild type virus (9, 10).

New York City (NYC) was among the first locations in Europe and North America to experience a large COVID-19 outbreak: by May 2020, the number of reported cases exceeded 150,000 (11). The first Gamma VOC case in NYC was identified on March 10^th^, 2021, in the middle of the second COVID-19 wave. The proportion of Gamma-attributed cases in NYC was growing but was outcompeted first by Iota (B.1.526 lineage) and Alpha variants (12), and then by Delta in summer 2021. Importantly, NYC, unlike many other locations in the US and globally, has a large-scale genomic surveillance system deployed by the New York City Public Health Laboratory (NYC PHL) along with the establishment of the NYC Pandemic Response Lab (PRL), which paved the way for 14,600 SARS-CoV-2 genomes to be sequenced by NYC PHL and PRL between 1 January to 1 May 2021.

Here we used a phylogeographic approach to investigate patterns of introduction and local circulation of the SARS-CoV-2 Gamma VOC in NYC in March-April 2021. Our analysis shows that nearly all circulating Gamma transmission clusters were introduced from other US locations and not from outside the US. The majority of these clusters were established in NYC long before the first Gamma case was first detected in the city, indicating that international travel restrictions are not an effective mitigation strategy even if they are in place before the variant emerges in the location of origin, and that even a comprehensive genomic surveillance system might not be able to identify variants as soon as they arrive to new locations.

## RESULTS

### Genetic Sequence Data and phylogeographic analysis identified 16 NYC gamma lineage clusters

As of June 5^th^, 2021, 15,967 unique Gamma genome sequences have been deposited in GISAID from around the world, including 116 from NYC public health surveillance. We reconstructed a maximum parsimony tree using these sequences and found that Gamma lineages circulating in NYC were dispersed throughout the phylogeny, indicating multiple independent introductions (Suppl. Fig.1).

To identify locally circulating NYC Gamma transmission clusters (clusters of >=2 sequences with an ancestral location in NYC) and infer the locations from which these clusters were introduced into the city (ancestral locations before NYC), we performed Bayesian phylogeographic analysis on a subset of Gamma sequences (N = 1,461) that we selected as “relevant” to NYC infections based on the Maximum Parsimony phylogeny (details in Methods). We identified 16 NYC transmission clusters, defined as clades with ancestral locations estimated to be in NYC (Fig. 1). NYC clusters’ sizes ranged between 2 and 108 sequences (median = 4) and included between 1 and 41 NYC sequences (median = 2).

**Figure 1.**
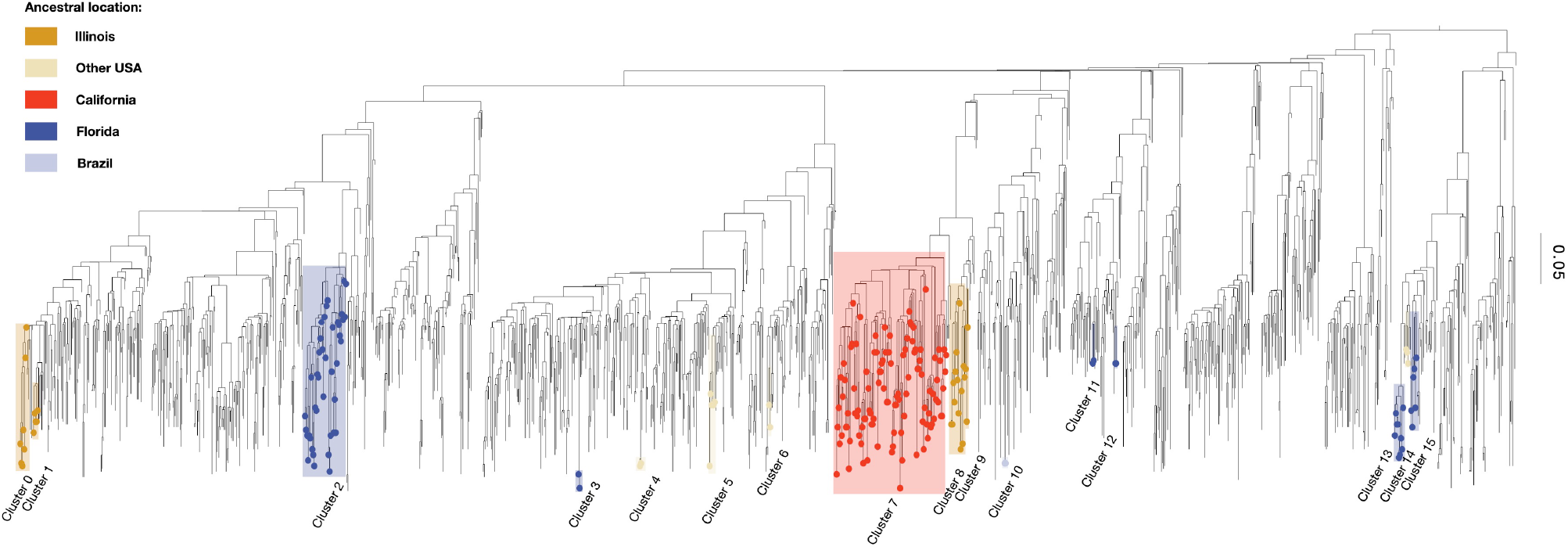
Molecular clock phylogenetic tree inferred from the subsampled Gamma lineages. Gamma clusters circulating in NYC in March and April 2021 are highlighted and color-coded by ancestral location.

The majority of NYC transmission clusters were introduced from Florida (N=6), followed by Illinois (N=4), and other locations in the US (N=4). Only one NYC Gamma transmission cluster (cluster 10, N = 2) was introduced from abroad, directly from Brazil (2). The two largest clusters, cluster 2 (N = 41) and cluster 7 (N = 108), included 16 and 41 NYC genomes and originated from the states of Florida and California, respectively (Fig. 1).

### The majority of Gamma lineages were persisting in NYC since early March 2021

Lineage persistence analysis identifies whether lineages have been introduced into the studied location within a defined time interval or they were already circulating in the given location before that time period (i.e. persisting lineages) (13). To describe patterns of introduction and persistence of Gamma lineages in NYC, we ran a lineage persistence analysis on the full Gamma dataset, as implemented in BEAST, focusing on the period between February 6^th^ (a month before the first Gamma lineage was detected in NYC) and May 1^st^ (the end of the second epidemic wave in the city). This analysis allowed us to describe whether local lineages had been circulating in NYC more than two weeks prior (persisting lineages) or if they were recently introduced from other locations.

In February 2021, less than 1% of NYC Gamma cases were associated with lineages imported from abroad. Gamma lineages were mainly introduced from “Other” US states (states that were not identified as separate discrete locations in our analyses, 59% of all introductions), followed by the introductions from New Jersey (16%), other parts of the New York state (9%), and Florida (8%), with the proportion of introductions from all other considered locations and local NYC persistence (lineages circulating in the city since 2 weeks prior) <3% each (Fig.2). By March 20^th^, 57% of all Gamma cases were persisting in NYC for at least two weeks; among the lineages that were newly introduced to NYC between the 6^th^ and 20^th^ of March 2021, the majority came from “Other” US states (22%), New Jersey (15%), other parts of New York State (4%) and Florida (2%). By the end of the second wave of the epidemic, in the second half of April 2021, 43% of Gamma lineages were persisting in NYC, 24% came from other parts of New York State, 9% from New Jersey, 3% from Florida, and 21% from “Other” US states. Though the majority of circulating NYC Gamma clusters identified through the phylogeographic analysis were introduced from Florida and Illinois, only a small proportion (around 5% and 1%, respectively) of Gamma lineages introduced in the observed period were from these states.

**Figure 2.**
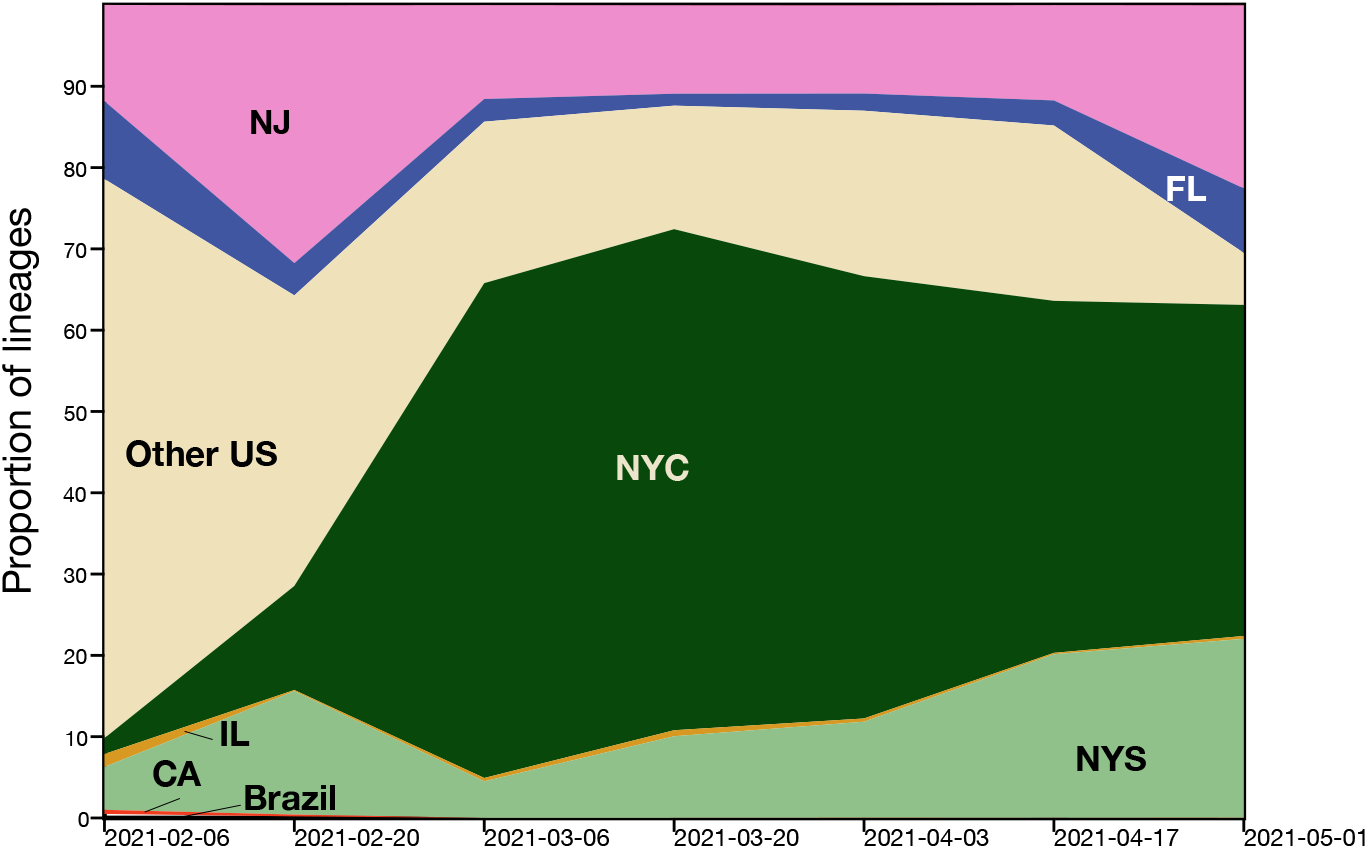
The proportion of Gamma lineages introduced (from New Jersey (NJ), Florida (FL), Illinois (IL), California (CA), New York State (excluding NYC; NYS), other domestic sources (Other US), and Brazil) and persisting (NYC) from various location in the US and abroad between February 6^th^ and May 1^st^ 2021. New York State (NYS). The NYC portion represents lineages persisting in NYC for more than two weeks since their introduction.

### Gamma transmission clusters were established in NYC in February 2021

Phylodynamic analysis of the times to most recent common ancestor (TMRCAs) for the identified Gamma transmission clusters showed that the majority of locally circulating clusters had been established in February 2021. The cluster TMRCAs ranged between January 5^th^ to May 18^th^, 2021 (Fig. 3). The TMRCAs for the largest circulating clusters, clusters 2 and 7, were February 19^th^, 2021 (95% highest posterior density [HPD] February 9 – 28, 2021) and February 25^th^, 2021 (95% HPD February 15^th^ – March 9^th^, 2021), respectively. The TMRCA for the only transmission cluster introduced from Brazil was estimated to be May 3^rd^, 2021 (95% HPD April 24^th^ and May 5^th^, 2021).

**Figure 3.**
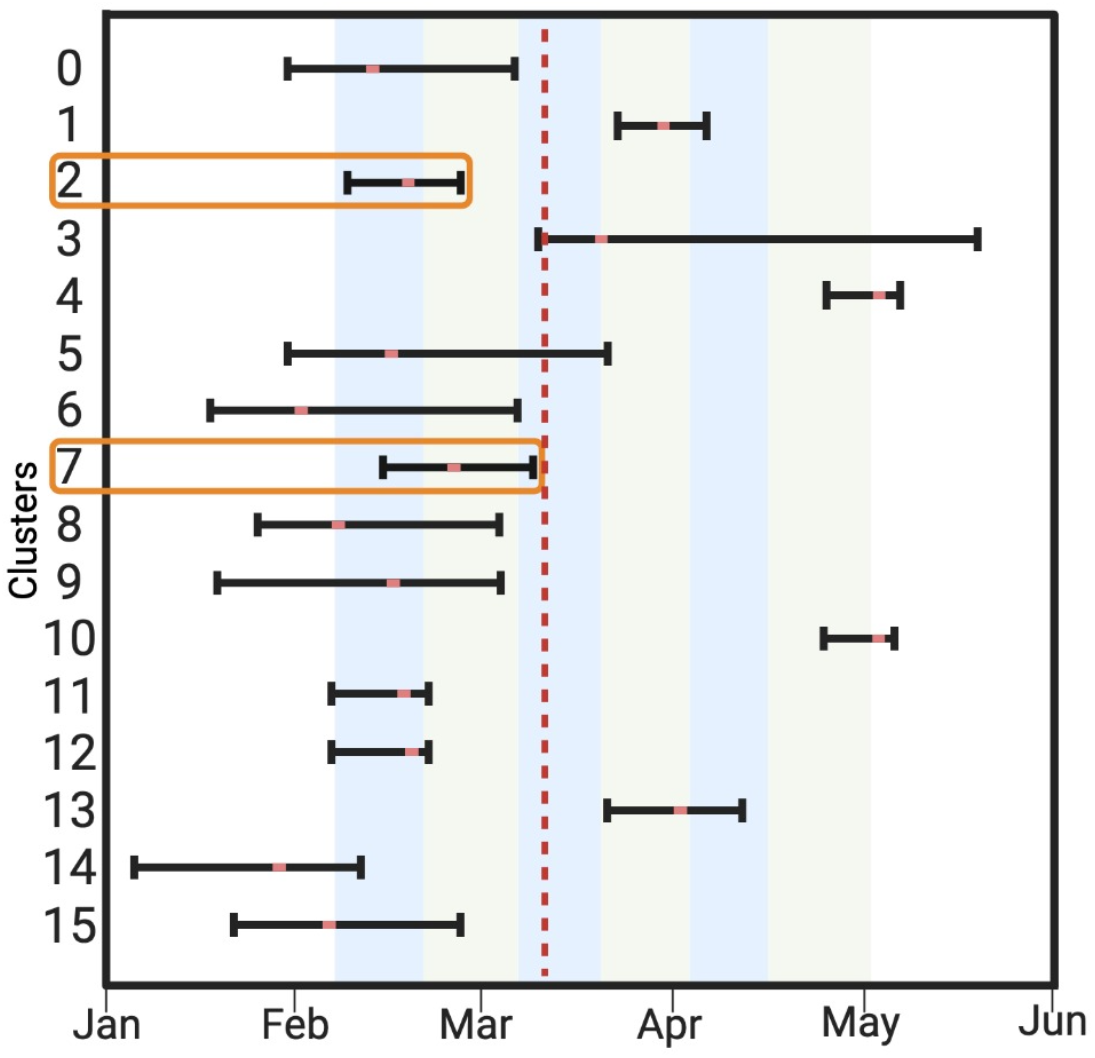
TMRCAs for Gamma transmission clusters circulating in NYC in early 2021. The red dotted line corresponds to March10^th^, when the first Gamma sequence was identified in NYC. The blue and green shaded areas correspond to the six two-week intervals in the analysis of persisting lineages. Orange squares highlight TMRCAs of the two largest clusters (N = 41 and N = 108 genomes).

### Gamma variant in neighborhoods with high pre-existing seropositivity in NYC in March – April 2021

Between March 6^th^ and May 1^st^, 2021, NYC PHL and PRL sequenced 11,385 SARS-CoV-2 genomes and identified 116 of them as Gamma using Pangolin (14). The two NYC neighborhoods (as defined by the United Hospital Fund) with the highest number of identified Gamma genomes were in Queens and Brooklyn (West Queens and East New York, Fig.4A). While the average proportion of SARS-CoV-2 sequences identified as Gamma remained low throughout city neighborhoods between March 6 and May 1 (0.7%, 95% CI 0.45 – 0.95 (15)), it was >2% in Chelsea, Greenpoint, Bayside Meadows, South East Queens, and East New York in that period. Some of these areas, including South East Queens and East New York, also showed some of the highest seropositivity rates observed in NYC as of March 11^th^, 2021 ((15), Fig. 4B), suggesting a more effective Gamma transmission in the presence of other circulating lineages in these areas with pre-existing immunity.

**Figure 4.**
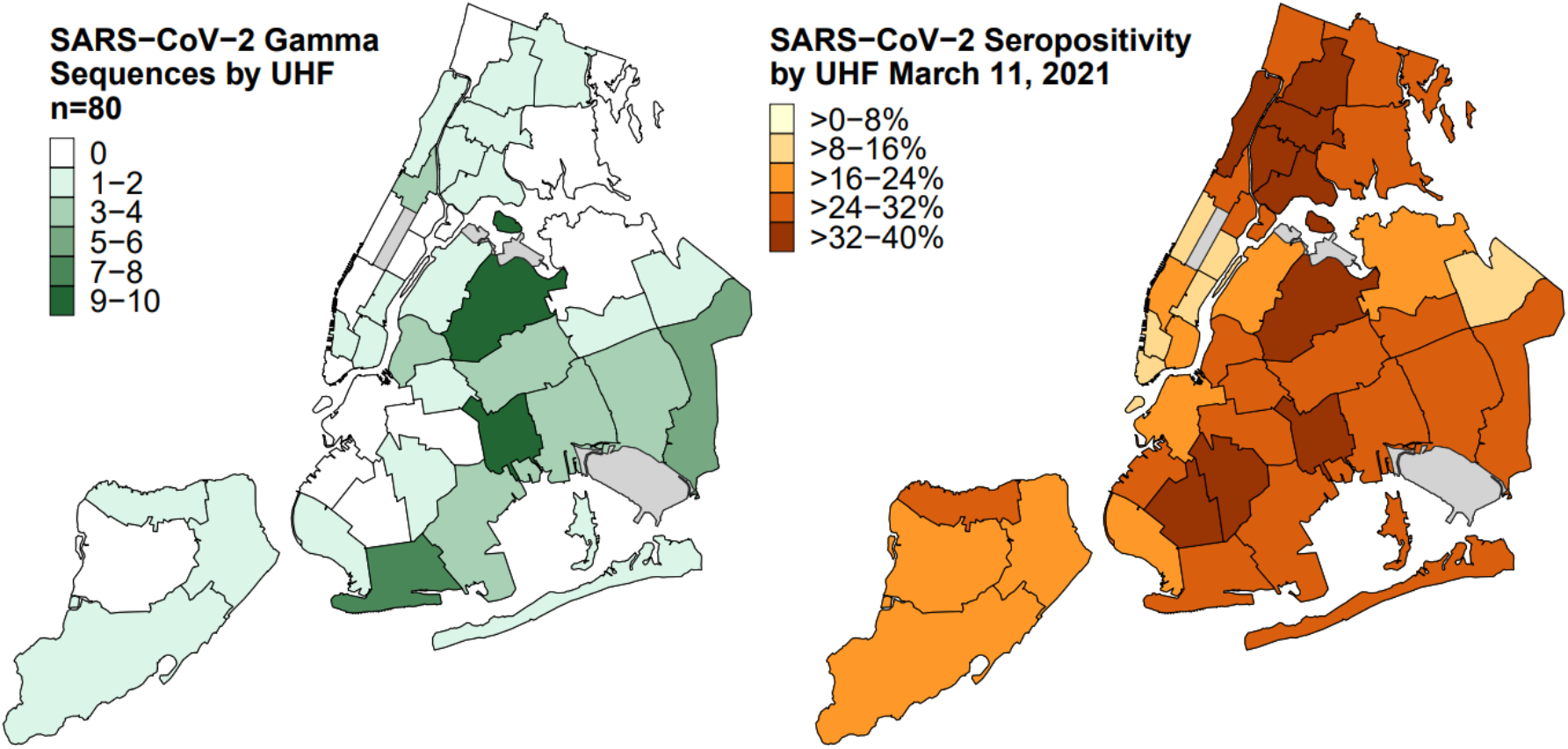
A) Geographical distribution of Gamma sequences sampled in NYC between March 10^th^ and May 1^st^, 2021. B) NYC city-wide seropositivity as of March 11^th^, 2021.

The first three Gamma lineages were found in Greenpoint, Bayside-Meadows, and Southeast Queens before March 19^th^, 2021 (Suppl. Fig.2). In the second half of March, at which point most Gamma lineages were already established locally and persisting in NYC, they became prevalent in West Queens and Coney Island – the areas where the majority of SARS-CoV-2 lineages were circulating in March-April 2021. In late April 2021, when the number of COVID-19 infections dropped in NYC, the Gamma lineages were most prevalent in Jamaica. Upon further investigation of the geographical spread of the Gamma lineages identified in the two largest circulating NYC clusters, clusters 2 and 7, no localized geographical patterns were observed: infections from both clusters were found throughout the city (Suppl. Fig.2).

## DISCUSSION

The emergence and swift spread of SARS-CoV-2 variants have threatened pandemic control efforts. To date, countries with well-developed SARS-CoV-2 genomic surveillance systems that have enabled early identification of newly emergent VOCs, such as South Africa and the United Kingdom, have been subjected to travel restrictions by other countries after alerting the international community of the new variant (4). Such measures might have substantial socio-political and economic consequences, but have not prevented the global dissemination of the variants and the subsequent infections growth (16). Here we show how the introduction and spread of the Gamma variant in NYC illustrates that although travel restrictions might reduce introductions from the origin locations, they failed to prevent variant establishment, as local NYC transmission clusters were seeded by domestic transmission.

Using a phylogeographic approach, we showed that despite the limited travel from Brazil - the country where Gamma lineage was first identified in late 2020 (2) - being enforced since May 2020, most Gamma lineages arrived in NYC and established locally circulating clusters in February 2021. All but one identified Gamma transmission clusters circulating in NYC were introduced from US locations, suggesting that the variant has been widely circulating in the US at the time and travel restrictions with Brazil did not prevent the variant from establishing in NYC. Only one of the Gamma transmission clusters circulating in NYC resulted from an introduction from outside of the US when the virus was imported from Brazil, but this introduction (in May 2021) was preceded by multiple introductions from domestic locations. Similarly, previous analyses of genomic, epidemiological, and travel data showed minimal evidence of direct SARS-CoV-2 transmission to NYC from China, the country where the virus was first identified, in early 2020; all other identified introductions that resulted in local NYC transmission were from other locations in the US (11). International travel restrictions imposed on China in early 2020 likely reduced the number of virus importation events but only modestly affected epidemic trajectory globally; their effect could have been greater if combined with local interventions that reduce transmission (17). Other evidence suggests that limiting travel might only have an impact on epidemic dynamics in countries with low COVID-19 incidence (18): the high daily number of COVID-19 cases in the US in early 2021 (19) might have contributed to a limited role of international introductions in the Gamma transmission. We show that once introduced, a large proportion of Gamma lineages persisted in NYC, also suggesting local mitigation strategies may have a much more substantial impact on variant spread than international travel restrictions.

As expected, in the presence of other dominant SARS-COV-2 strains such as the Iota and the Alpha variants, Gamma spread more effectively in areas with high seroprevalence than in other neighborhoods: NYC areas with some of the highest seroprevalence levels were also those with the highest number of Gamma cases. Vaccine rollout undoubtedly slowed down the spread of Delta (20), but its impact on Gamma spread in NYC was unlikely as pronounced. Even though vaccination rates were accelerating in early 2021 (the number of people who received at least one dose of the vaccines grew from 7% on February 1^st^ to 18.8% on March 10^th^ to 45.4% on May 1^st^ (15)), these efforts were delayed relative to the time of Gamma introduction and establishment in the city. Thus, the most successful Gamma lineages that resulted in large locally-circulating clusters were widespread in the city and were not constrained to specific neighborhoods.

NYC dramatically increased its sequencing capacity in January-February 2021 with the implementation of sequencing at PRL: in February 2021, in the middle of the second epidemic wave, 4-6% of all SARS-CoV-2 infections in NYC were genotyped (15), which is much higher than average (1.75%) in North America (1). Although these efforts allowed for timely identification of the rapidly-growing Iota variant that originated in NYC (12), they did not help to substantially reduce a lag between the introduction and identification of Gamma lineages, which were present at much lower frequencies, in NYC. This lag was similar to the one recorded in the early days of the pandemic in the absence of available widespread testing when the first SARS-CoV-2 case in NYC was registered on February 29^th^, 2020 but phylodynamic analysis later revealed that the virus has likely been introduced to NYC as early as January (11, 21). Importantly, laboratory surveillance includes a necessary lag from the sample collection to laboratory analysis and quality checks before a newly emerging variant can be reported. In NYC, the public health laboratory is continuously improving systems and processes to minimize this lag and encourages timely reporting of genomic data by hospitals and academic laboratories that led to substantial improvement in volume and timeliness of genomic data in each subsequent SARS-CoV-2 pandemic wave.

Patterns of introduction from within the US showed that a small number of introductions can result in a large number of locally circulating clusters. Even though <5% of the circulating NYC Gamma lineages, which includes circulating Gamma clusters and singletons, were introduced from the state of Florida in February-April 2021, introductions from Florida seeded the majority of circulating NYC Gamma transmission clusters. Similarly, even though the overall proportion of Gamma lineages introduced from Illinois was <1% at all times during the observation period, these introductions seeded 4 of the 16 (25%) identified circulating transmission clusters. These patterns are similar to those previously reported for Alpha VOC (B.1.1.7): the lineage first identified in the UK was initially introduced into three locations in the US (California, Florida, and Georgia) and then spread through unmitigated transmission between different US locations, with Florida state playing an important role in that transmission (22).

Similar to any such analysis, our work is affected by the disproportional sampling and sequencing efforts in various US locations and abroad. As our analysis only focused on the introductions and establishment of Gamma into NYC, we did not investigate importations of Gamma into other domestic locations. Furthermore, our subsampling approach was based on maximum parsimony phylogenetic reconstruction which could have affected the representation of different geographic locations in our final dataset but was necessary to create a dataset that could be analyzed in a Bayesian framework. Both the development of methods that can analyze larger datasets to avoid down-sampling, and further improvement in genomic surveillance capacity proportionally across the US locations and worldwide can help improve similar investigations in the future.

The analysis of the introduction and spread of Gamma lineages in NYC, a location with large-scale genomic surveillance, shows that even in the presence of effective genomic monitoring efforts, low-prevalence variants with potential public health importance can circulate undetected amidst an ongoing epidemic. Even though US travel restrictions might be effective at reducing the number of direct introductions from the international locations where the variant was first identified, they do not preclude the establishment of local transmission clusters if the variant has already established itself elsewhere domestically.

## METHODS

### Sequence data

To characterize the emergence of the Gamma variant in NYC, we downloaded all complete high-quality (<5% ambiguous sites) with an available date of sampling (day, month, year) SARS-CoV-2 Gamma genomes available through the GISAID database from March 10^th^ to June 5^th^ as of June 5^th^ (N = 15,967) (23). After removing duplicate sequences from all locations except for the location of interest (NYC), we aligned the remaining Gamma sequences using MAFFTv.7.453 (24), trimmed the 5’ and 3’ ends and masked all problematic sites in the alignment as suggested before (25). IRB of UC San Diego waived ethical approval of this work as human subjects except.

### Phylogenetic analysis and downsampling

To identify phylogenetic clusters comprising of lineages predominantly circulating in NYC, we first inferred a maximum parsimony (MP) phylogenetic tree representing the Gamma global diversity using all available unique sequences and IQTreev.2.0.3 software (26). We then removed all clades that did not have NYC sequences and were not ancestral to clades that contained NYC sequences. For large polytomies, a random sub-sampling of genomes was performed. The resulting subsampled dataset was further used to reconstruct a maximum likelihood (ML) phylogenetic tree under the GTR+F+I nucleotide substitution model, collapsing polytomies, and enforcing a minimum branch length of 1e-9.

### Phylogeographic analysis

We performed Bayesian phylogeographic reconstruction in BEAST1.10.4 (27). First, we reconstructed time-scaled phylogenetic trees based on the genomes in the subsampled dataset under an HKY85 nucleotide substitution model, assuming a strict molecular clock model (fixed to the value of 8×10^−4^ nt/site/year) and a Bayesian skyline coalescent tree prior (28). We ran 3 independent MCMC chains of 1×10^9^ generations; for all BEAST analyses, we used Tracer (29) to assess convergence of MCMC chains and to ensure ESS>100 for all parameters. We used the LogCombiner package from BEAST to combine the tree distributions from these three independent chains and resample them at a lower frequency to obtain a distribution of 1,000 trees. The phylogeographic reconstruction of viral lineage movement was then performed using the resulting tree distribution and assigning geographic trait value to the tips of the phylogeny based on the sampling locations. Phylogeographic analysis assumes that location changes follow the same process as sequence evolution; we used an asymmetric model that allows estimating separate incoming and outgoing viral lineage flow rates for each location. We considered 10 locations in our analysis: (1) NYC, (2) Other New York locations, (3) California, (4) Florida, (5) Illinois, (6) New Jersey, (7) Other USA locations, (8) Brazil, (9) Other South America Locations, (10) Other locations in the world. The specific states were selected based on the high number of Gamma lineages reported. Brazil was selected as the country where the Gamma VOC was first identified (2); similarly, other South American countries were included as a separate location because of the close proximity to Brazil and the high number of reported Gamma cases. We then ran another MCMC analysis of 5×10^6^ generations which allowed us to obtain posterior tree distributions annotated with Markov jumps indicating virus migration events between the specified geographical locations.

Gamma clades circulating in NYC were then identified in a phylogeographic context, defined as clades of ≥2 sequences for which the ancestral location was in NYC with 90% posterior probability support. The ancestral location of NYC clades was then identified as the ancestral location of the ancestral node that had the highest posterior probability support.

### Analysis of persisting lineages

To describe the proportion of circulating Gamma strains versus those that were being introduced from other locations, we ran lineage persistence analysis using PersistenceSummarizer as described by Lemey et al (13). This analysis allowed us to describe the proportion of lineages being introduced to NYC from other locations within each 2-week period and the proportion of lineages that continued circulating in NYC from before the start of the period (those being “re-introduced” from within NYC). We split the time into 2-week intervals going backward from the most recently sampled sequence (June 5^th^, 2021). Since the first Gamma sequence was identified in NYC on March 10^th^, 2021, we analyzed the introduction and persistence of circulating lineages for six 2-week time periods starting two weeks before the first week with a non-zero number of Gamma sequences (February 6^th^) and until the end of the second epidemic wave in NYC (May 1^st^).

## Data Availability

All data used in the analyses are available though GISAID (https://www.gisaid.org/).

## Acknowledgments

T.I.V. is funded by a Branco Weiss Fellowship. J.O.W. acknowledges funding from the National Institutes of Health (AI135992). G.W.H. acknowledges funding from the National Institutes of Health (AI154824). K.G.A acknowledges funding from the National Institutes of Health (U01AI151812 and U19AI135995) and CDC BAA contract 75D30120C09795. This work was supported (in part) by the Epidemiology and Laboratory Capacity (ELC) for Infectious Diseases Cooperative Agreement (Grant Number: ELC DETECT (6NU50CK000517-01-07) funded by the Centers for Disease Control and Prevention (CDC). The findings and conclusions in this report are those of the authors and do not necessarily represent the official position of the Centers for Disease Control and Prevention. Use of trade names is for identification only and does not imply endorsement by the Centers for Disease Control and Prevention. We thank Mustapha Mustapha of the New York City Department of Health and Mental Hygiene for his useful comments on the manuscript text.

## FIGURES

**Supplementary Figure 1.**
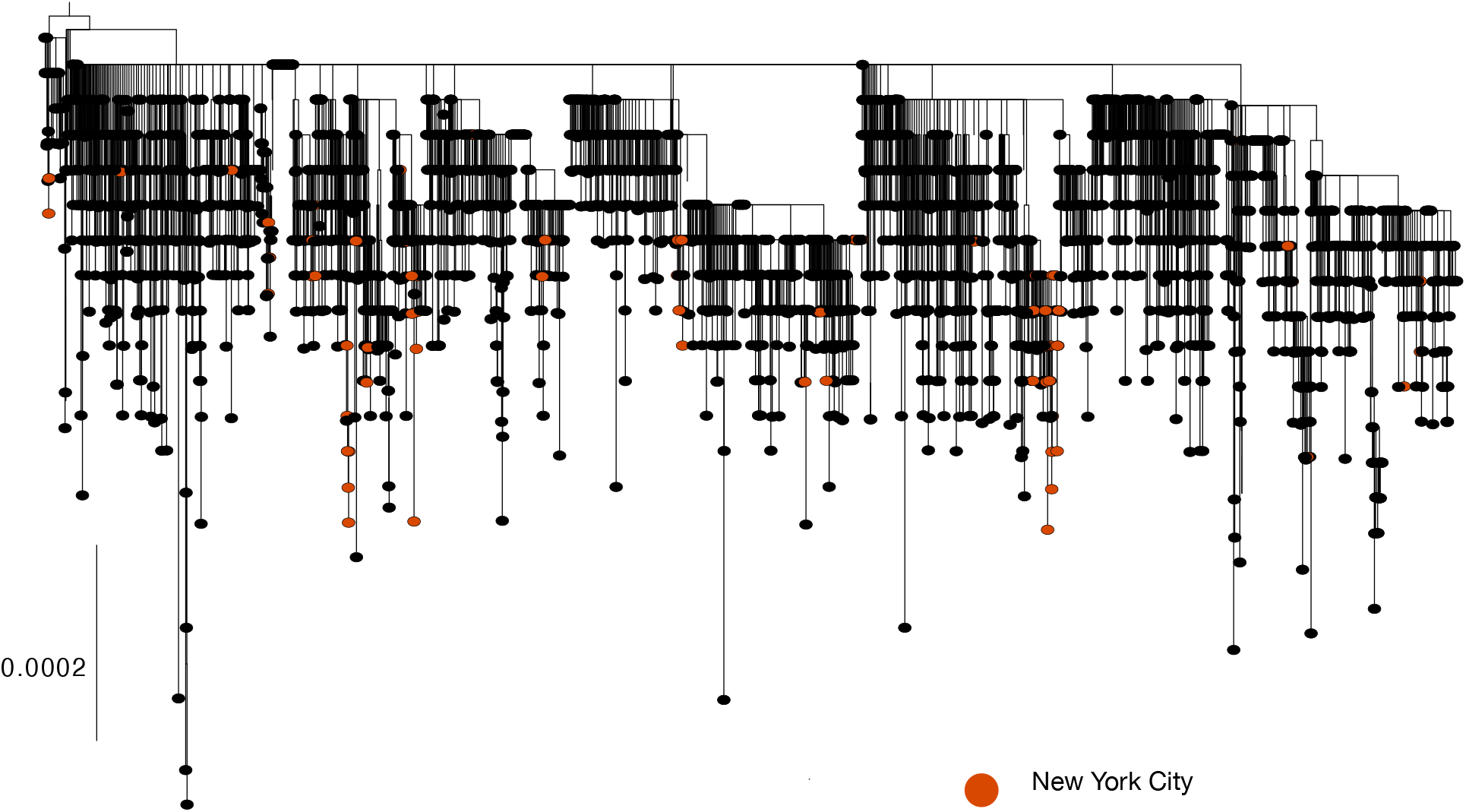
Global maximum parsimony phylogeny of the full set of SARS-COV-2 Gamma lineages circulating between March 10^th^ and June 5^th^, 2021.

**Supplementary Figure 2.**
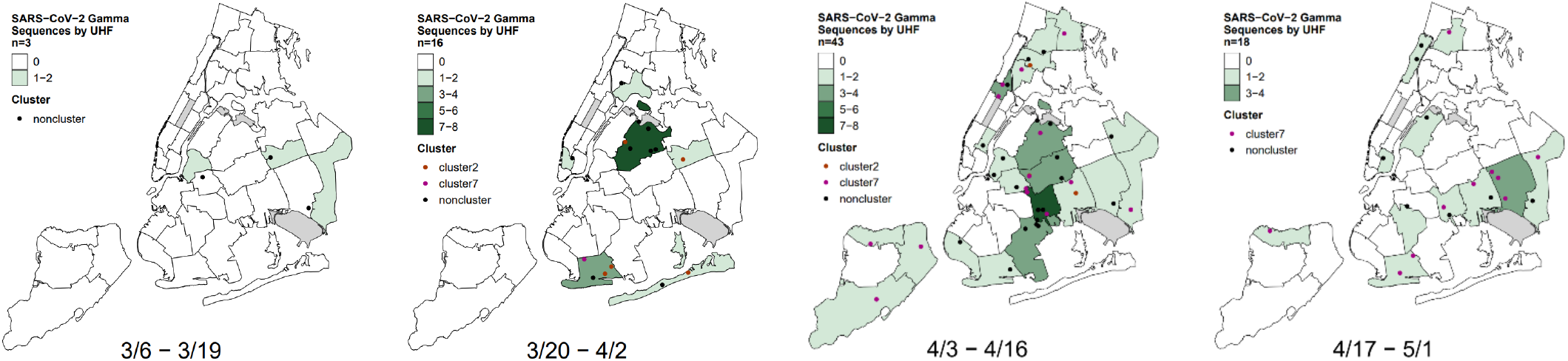
Changes in the geographical distribution of SARS-CoV-2 Gamma variant in NYC over time.

